# Epidemiological characteristics of COVID-19 in medical staff members of neurosurgery departments in Hubei province: A multicentre descriptive study

**DOI:** 10.1101/2020.04.20.20064899

**Authors:** Qiangping Wang, Xing Huang, Yansen Bai, Xuan Wang, Haijun Wang, Xuebin Hu, Feng Wang, Xianke Wang, Jincao Chen, Qianxue Chen, Xiaobing Jiang, Hongyang Zhao

## Abstract

**Background:** The novel coronavirus (SARS-CoV-2) has infected a large number of healthcare workers in Hubei province, China. In addition to infectious and respiratory disease physicians, many doctors in other medical fields have been infected.

**Methods:** We prospectively collected epidemiological data on medical staff members who are working in neurosurgery departments in 107 hospitals in Hubei province through self-reported questionnaires or telephone interviews. Data of medical staff members with laboratory-confirmed coronavirus disease 2019 (COVID-19) were analysed. The final follow-up date was 1 March 2020.

**Findings:** A total of 5,442 neurosurgery department medical staff members were surveyed. One hundred and twenty cases, involving 54 doctors and 66 nurses, were found to have been infected with SARS-CoV-2. The overall incidence was 2.2%. These cases were concentrated in 26 centres, 16 of which had admitted a total of 59 patients with COVID-19 complicated by craniocerebral disease. Medical staff members in centres receiving COVID-19 patients had a higher risk of contracting infection than those in centres not receiving COVID-19 patients (relative risk: 19.6; 95% confidence interval: 12.6–30.6). Contact with either COVID-19 patients (62.5%, 75/120) or infected colleagues (30.8%, 37/120) was the most common mode of transmission. About 78.3% (94/120) of the infected cases wore surgical masks, whereas 20.8% (25/120) failed to use protection when exposed to the source of infection. Severe infections were observed in 11.7% (14/120) of the cases, with one death (0.8%, 1/120). All the infected medical staff members had been discharged from the hospital. A total of 1,287 medical staff members were dispatched to participate in the frontline response to COVID-19 under level 2 protection of whom one was infected. Medical staff members who took inadequate protection had a higher risk of contracting infection than those using level 2 protection (relative risk: 36.9; 95% confidence interval: 5.2–263.6).

**Conclusions:** Neurosurgical staff members in Hubei province were seriously affected by COVID-19. Level 2 protection and strengthening of protective measures are likely to be effective in preventing medical workers from being infected.

## Introduction

Since December 2019, Wuhan, in the Hubei province of China, has become a global focus of attention due to an outbreak of viral pneumonia.^1^ The pathogen involved has been confirmed as a novel coronavirus, which has been named ‘severe acute respiratory syndrome coronavirus 2’ (SARS-CoV-2).^1^ The SARS-CoV-2 causes an acute respiratory illness known as ‘COVID-19’ (Corona Virus Disease 19).^2-6^

SARS-CoV-2 is phylogenetically similar to SARS-CoV and has been considered as a sister of the SARS-CoA virus, belonging to the species of severe acute respiratory syndrome-related coronaviruses. ^7 8^ COVID-19 patients exhibit symptoms similar to those of SARS patients, such as fever, non-productive cough, dyspnoea, fatigue and radiographic evidence of pneumonia.^9^ Human-to-human transmission has been confirmed as a significant factor for the development of both COVID-19 and SARS, resulting in a rapid spread of the disease. ^10-13^ According to the reports on the COVID-19 situation, which were released by the World Health Organisation, as of 3 April 2020, China had reported a total of 82,802 cases of COVID-19, including 3,331 associated deaths. Around the world, there are 972,303 cases in 49 countries, with 50,322 fatalities. The disease is already taking a far more extensive toll on global public health than SARS did in 2002; SARS caused more than 8,000 infections and 700 deaths.^14^

Due to exposure to SARS-CoV-2-infected patients, healthcare workers are likely to be victims of this infectious disease.^15^ As of 11 February 2020, more than 1,700 healthcare workers had been infected in China, including more than 1,400 in Hubei province.^16^ Recently, the clinical characteristics of COVID-19 patients were revealed through large-scale population studies. ^9 13 17 18^ These have provided a comprehensive insight into the new disease, which may lead to better prevention and treatment. However, reports on the epidemiological and clinical features of COVID-19 in infected medical staff members are still limited. Not only infectious and respiratory disease physicians but also many doctors in other medical fields have been infected. In Hubei province, more than 10 medical staff members in neurosurgery departments were found to be infected in each of four hospitals. In this study, we investigated 107 hospitals in Hubei province, aiming to unravel the epidemiological characteristics of COVID-19 in the medical staff members of the neurosurgery departments affected by SARS-CoV-2 in this region.

## Methods

### Study design and participants

We conducted a retrospective study focusing on the epidemiological characteristics of medical staff members in neurosurgery departments in Hubei province who were affected by SARS-CoV-2. The study was supported by the Neurosurgical Branch of Hubei Medical Association and approved by the institutional ethics board of Union Hospital, Tongji Medical College, Huazhong University of Science and Technology, Wuhan, China (No. 20200029). Patients or members of the public were not involved in the design, conduct, reporting or dissemination of the research. We first sent out questionnaires to the directors of the member units of the Neurosurgical Branch of Hubei Medical Association to obtain department demographic data and basic information about infected medical staff. We conducted epidemiological investigations on infected medical staff members through self-reported questionnaires or telephone interviews with the assistance of directors.

All COVID-19 medical staff members in the neurosurgery departments affected by COVID-19 were enrolled and tested according to the *Sixth Edition of the Diagnosis and Treatment of Novel Coronavirus Pneumonia*, published by the office of the National Health Commission of the People’s Republic of China (http://www.nhc.gov.cn/). Due to the uncertainty of clinically confirmed cases, only participants with a positive nucleic acid test result on real-time reverse-transcriptase–polymerase chain reaction (RT-PCR) assay of nasal and pharyngeal swab specimens were considered cases. The detection method is the same as that described in detail by Liu et al. ^15^

The infection source was comprehensively evaluated based on contact time, the protective measures when exposure occurred and onset time. The earliest patient who had been in close contact (within 1 m) while taking the weakest precautions was presumed to be the most likely source of infection. The protection standards for infectious diseases in China was graded into three levels according to the following criteria: Level 1 protection: white coat, disposable hat, disposable isolation clothing, disposable gloves and disposable surgical mask (replace them every 4 h or when they are wet or contaminated); Level 2 protection: disposable hat, medical protective mask (N95 or higher standard), goggles (anti-fog) or protective mask (anti-fog), medical protective clothing or white coats covered by medical protective clothing, disposable gloves and disposable shoe covers; Level 3 protection: as for level 2 protection, replacing the goggle (anti-fog) or protective mask with a comprehensive respirator or higher-level respirator with an electric air supply filter (positive pressure head cover).

The severity of COVID-19 (severe *vs*. non-severe) was defined according to the American Thoracic Society guidelines for community-acquired pneumonia.^19^ The incubation period was defined as the interval between the earliest date of contact with the transmission source and the earliest date of onset of symptoms. The date of onset of the disease was defined as the day when one or more symptoms were noticed. The discharge criteria were as follows: (1) the body temperature had returned to normal for more than 3 days, and the respiratory symptoms had significantly improved; (2) lung imaging revealed obvious reduction in inflammatory changes; (3) two consecutive negative nucleic acid tests for respiratory pathogens, with a sampling interval of at least 1 day. Those who were able to meet these criteria were considered as recovered. The final follow-up date was 1 March 2020.

### Data collection

Epidemiological and clinical characteristics were obtained using data collection forms administered through questionnaires and telephone-based follow-up interviews. The data were reviewed by a trained team of physicians and statisticians. The general data for each single centre included hospital grade, the number of medical staff members and infected cases, COVID-19 patients admitted and the number of staff members allocated to the management of COVID-19 patients. Information on infected staff members included data on sex, age, exposure history, protective measures when exposure occurred, incubation period, onset of infection, time of confirmed infection, disease severity and prognosis.

### Data analysis

All statistical analyses were conducted using the SPSS (Statistical Package for the Social Sciences) version 19.0 software (SPSS, Chicago, IL, USA). Continuous variables were described using mean, median and distribution range values. Categorical variables were expressed as frequency rates and percentages. For the continuous variables of two independent samples, if the normal distribution was satisfied, the t-test was used, whereas if the normal distribution was not satisfied, the Mann–Whitney U test was employed. For categorical variables, chi-squared and Fisher’s exact tests were recommended. Relative risk and 95% confidence intervals were used to identify the relationship between the two different groups. After correction and comparison, a value of p < 0.05 was considered to be statistically significant.

## Results

### General information

Under the coordination of the Neurosurgical Branch of Hubei Medical Association, we obtained survey information from neurosurgery departments in 107 hospitals across 13 cities in Hubei province, China. A further nine hospitals were not included in the study because their directors did not respond. To protect privacy, the hospital names were hidden in this article. All living infected medical staff members responded to this study. Information on the doctor who died was obtained from his family members and colleagues.

According to the classification of hospitals in China, 51 (47.7%) third-grade class A, 25 third-grade class B (23.4%) and 31 (28.9%) second-grade class A hospitals were enrolled in this survey. The distribution of hospitals is presented in Figure 1. From these centres, 5,442 medical staff members comprising 1,757 doctors and 3,685 nurses were included. A total of 1,287 of these had been allocated to frontline management of COVID-19 patients since 20 January 2020.

**Figure 1.**
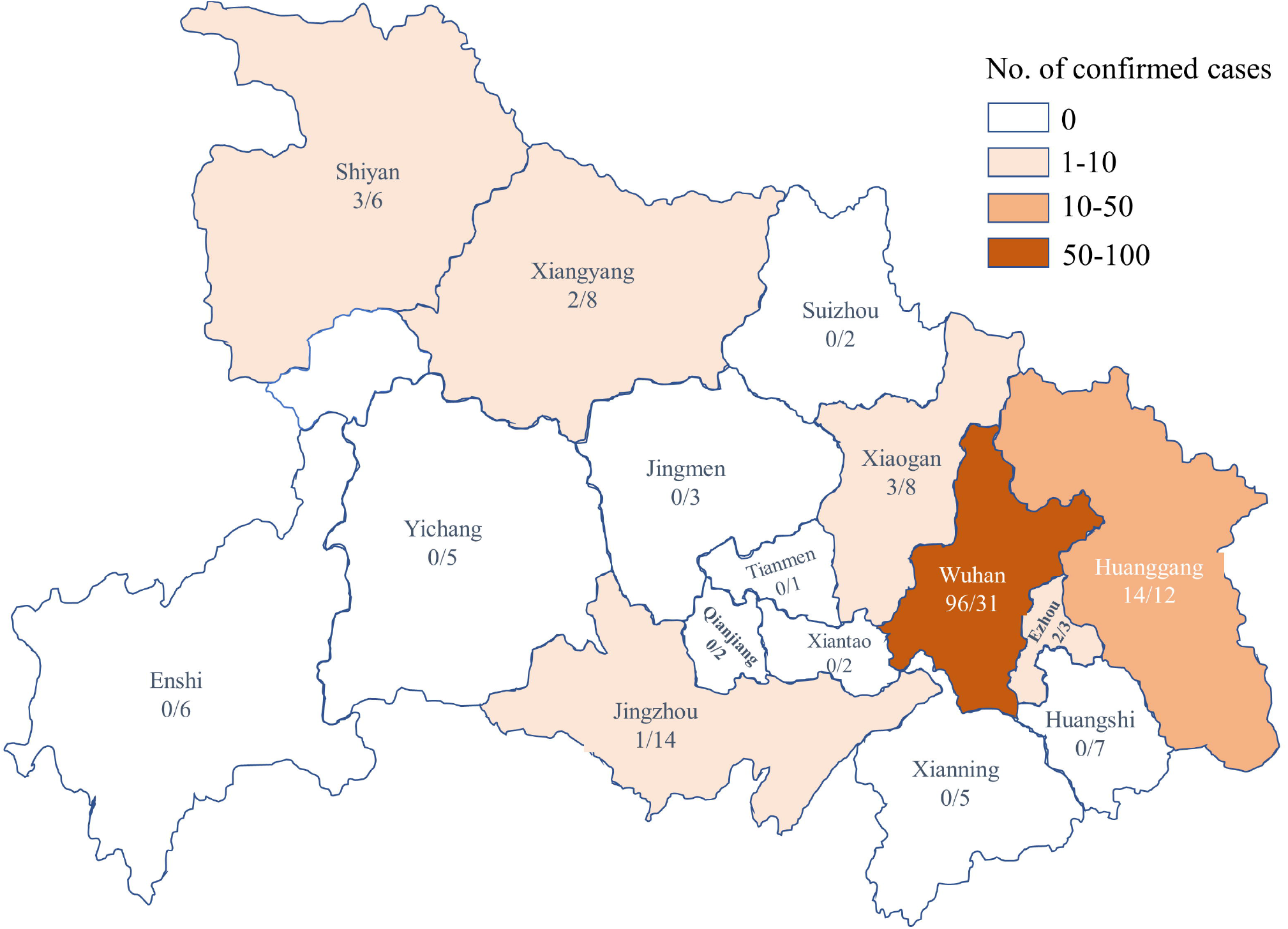
Distribution of centres and infected healthcare workers in neurosurgery departments across Hubei province, China. The numerator represents the number of infected healthcare workers, whereas the denominator represents the number of single centres.

There were 16 hospitals that have treated a total of 59 patients with craniocerebral disease who were subsequently diagnosed with laboratory-confirmed COVID-19. A centre in Huanggang had admitted up to 16 COVID-19 patients, and centres in Wuhan admitted 33 patients (56.0%). The number of COVID-19 patients admitted to each centre in Wuhan is presented in Figure 2B.

**Figure 2.**
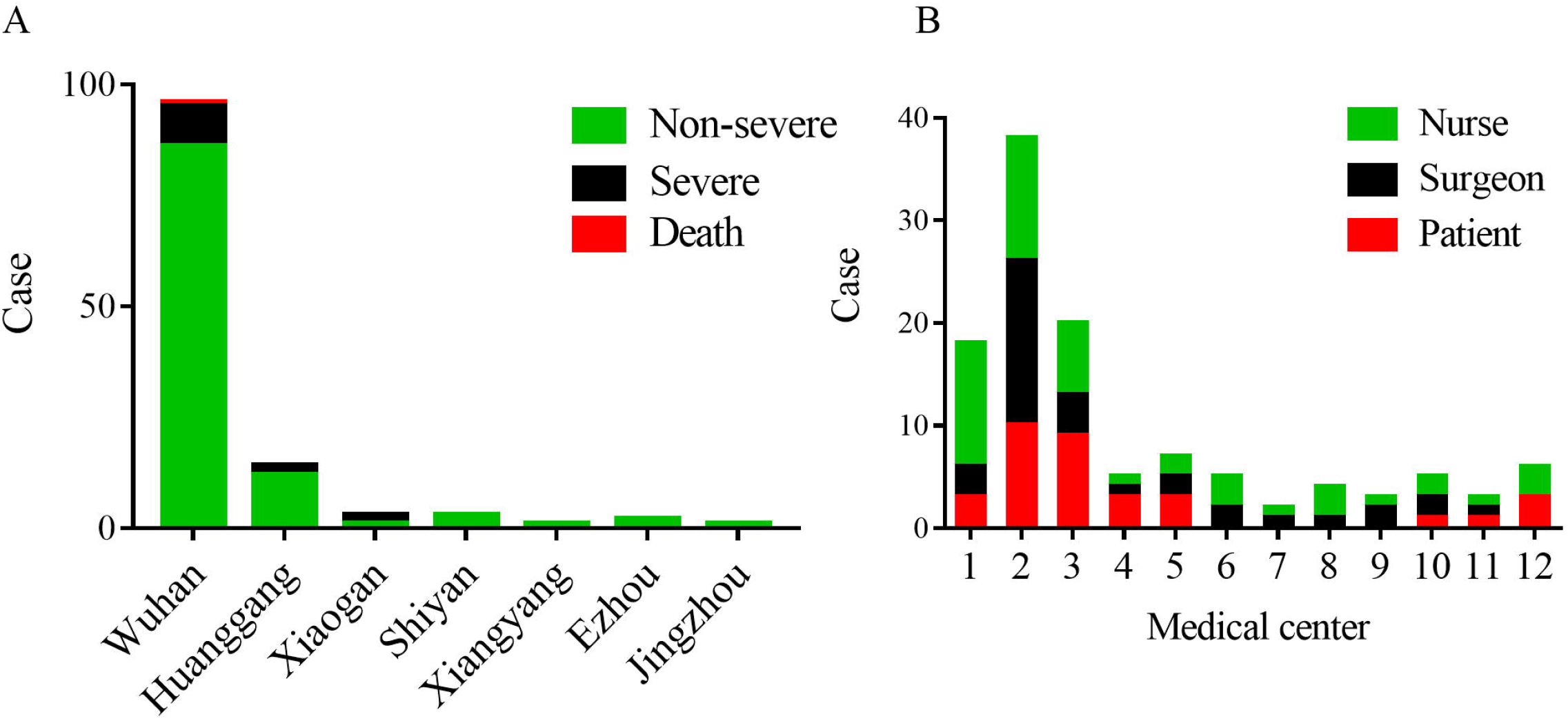
Distribution of infected healthcare workers and COVID-19 patients admitted to medical centres. A) Distribution of cases in seven cities. B) Distribution of infected healthcare workers and COVID-19 patients admitted to severely affected centres in Wuhan.

A total of 120 staff members in neurosurgery departments of 26 hospitals were infected with SARS-CoV-2, as confirmed by a nucleic acid test. The overall incidence was 2.2%, and 96 of the cases were in Wuhan (79.3%, Figures 1 and 2A). More than 10 staff members were infected in each of four centres, with a maximum of 28 in a centre in Wuhan. The distribution of infected cases is presented in Figure 1 and Figure 2B.

### Epidemiological characteristics of infected medical staff

There were 54 neurosurgeons and 66 nurses who were infected with SARS-CoV-2, including 60 males (50.0%) and 60 females (50.0%). The median age was 33.5 years (range, 23–51 years). The first occurrence of COVID-19 in this population was reported on 8 January 2020, with the number of confirmed cases reaching its peak on 19 and 29 January. The last case was confirmed on 9 February 2020. The data distribution regarding the date of onset of illness and diagnosis of the infected cases is presented in Figure 3A. The incubation period ranged from 1 to 11 days, with a median of 5.2 (±1.8) days (Figure 3B). There were 15 staff members (12.4%) who were found to be having severe disease and 106 having non-severe disease (88.3%). A total of 117 (97.5%) COVID-19-infected individuals were admitted to hospital, and only 3 (2.5%) non-severe cases were subject to home quarantine. Of the 120 COVID-19-infected medical staff, 1 died, and the remaining 119 patients (99.2%) had been discharged from hospital by 26 February 2020. No distinct complications were observed in the discharged patients. The demographic and epidemiological details of the infected medical staff members are summarised in Table 1.

**Table 1.** Characteristics of COVID-19 in medical staff members in Hubei province

|  | Total | Group 1 | Group 2 | P value |
| --- | --- | --- | --- | --- |
| Hospital number | 107 | 16(15.0%) | 91(85.0%) |  |
| Total staff | 5442 | 921(16.9%) | 4521(83.1%) |  |
| Infected staff | 120 | 96(80.0%) | 24(20.0%) | <0.001 |
| Gender |  |  |  | 1.000 |
| Male | 60(50.0%) | 48(50.0%) | 12(50.0%) |  |
| Femal | 60(50.0%) | 48(50.0%) | 12(50.0%) |  |
| Age | 33.5(23-56) | 34(23-56) | 31.5(25-51) | 0.820 |
| Occupation |  |  |  | 0.650 |
| Surgeon | 54(45.0%) | 42(43.8%) | 12(50.0%) |  |
| Nurse | 66(55.0%) | 54(56.2%) | 12(50.0%) |  |
| Protective measure when exposure |  |  |  |  |
| None | 25(20.8%) | 21(21.9%) | 4(16.7%) | 0.780 |
| Surgical mask | 94(78.3%) | 74(77.1%) | 20(83.3%) | 0.591 |
| Level-2 protection | 1(0.8%) | 1(1.0%) | 0 | 1.000 |
| Mode of infection |  |  |  |  |
| From patient | 75(62.5%) | 68(70.8%) | 7(29.2%) | <0.001 |
| From Colleague | 37(30.8%) | 28(29.2%) | 9(37.5%) | 0.464 |
| From family | 1(0.8%) | 0 | 1(4.1%) | 0.200 |
| Uncertain | 7(5.8%) | 0 | 7(29.2%) | <0.001 |
| Incubation period | 5.2±1.8(1-11) | 5.0±1.7(1-9) | 5.7±1.8(1-11) | 0.362 |
| Illness severity |  |  |  | 1.000 |
| Non-severe | 106(88.3%) | 85(88.5%) | 21(87.5%) |  |
| Severe | 14(11.7%) | 11(11.5%) | 3(12.5%) |  |
| Treatment |  |  |  | 0.491 |
| Home quarantine | 3(2.5%) | 2(2.1%) | 1(4.2%) |  |
| Hospitalization | 117(97.5%) | 94(97.9%) | 23(95.8%) |  |
| Clinical outcome |  |  |  | 0.200 |
| Recovery | 119(99.2%) | 96(100%) | 23(95.8%) |  |
| Death | 1(0.8%) | 0 | 1(4.2%) |  |

**Figure 3.**
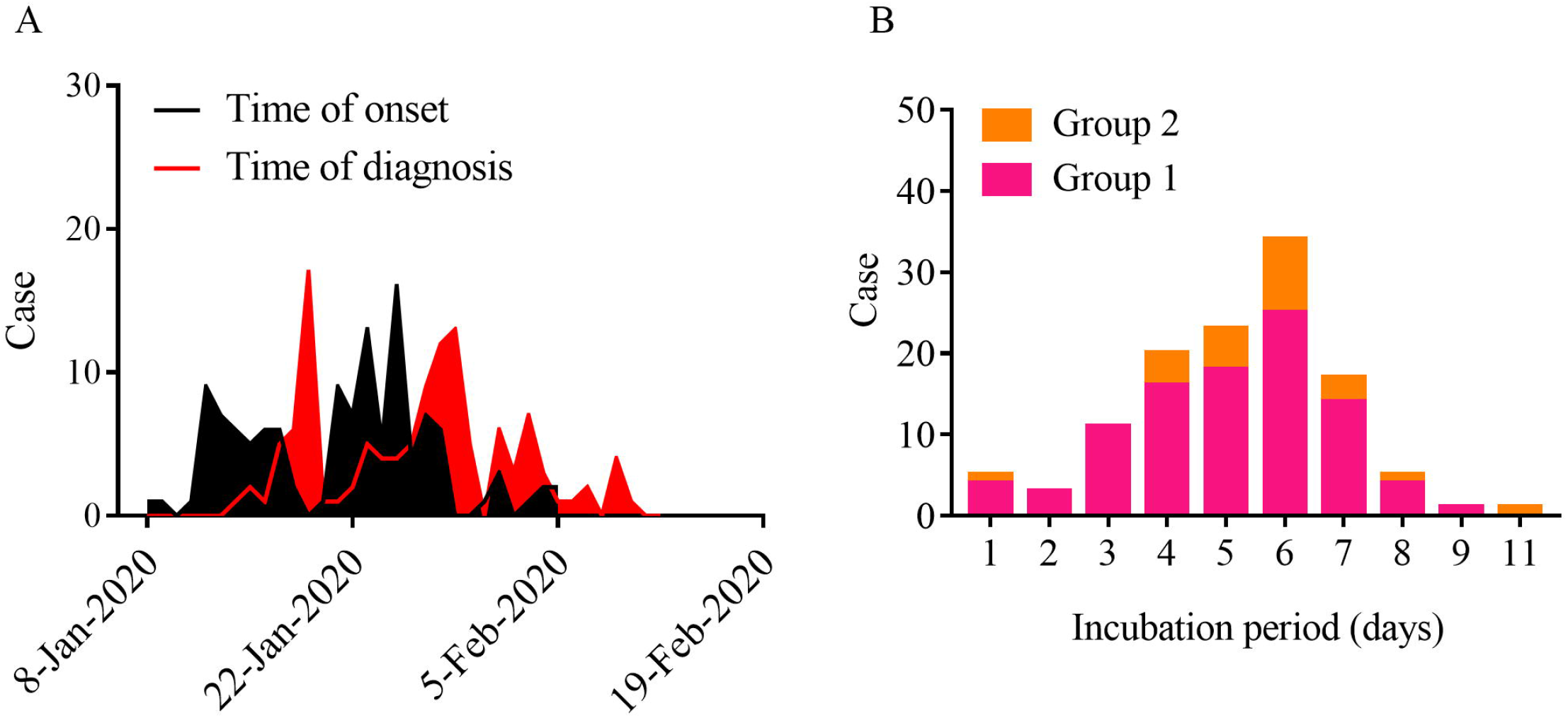
Distribution of the date of onset of illness and diagnosis (A) and the latent period (B). Group 1: the centres caring for craniocerebral patients with COVID-19 infection; Group 2: the centres not caring for craniocerebral patients with COVID-19 infection.

All of the 120 infected medical staff members participated in clinical work before the onset of their illness, and all of them had a history of direct contact with COVID-19 patients at a close distance (within 1 m), with the contact times ranging from 1 to 25 min, the average contact times 10 (4–17), the contact time ranging from 5 to 90 min, and the average cumulative contact time 35.4 min. Of the 120 infected staff, 119 did not use standard protective measures at work before infection, which means that only one was infected during the period under level 2 protection.

Seventy-five cases (62.5%) were most likely infected by direct contact with COVID-19-confirmed patients, whereas infections in 30.8% (37/120) of the medical staff members occurred upon contact with COVID-19-confirmed colleagues. One case was infected by a family member, and for seven cases, the infection source could not be identified with certainty because they could not recall the exact time of contact with more than one patient or infected colleagues. No confirmed cases of transmission from medical staff members to patients were noted. Ninety-four medical personnel (78.3%, 89/120) used a surgical mask, whereas 25 (20.8%) did not take any protective measures when exposed to the source of infection. Only 1 out of the 1,287 dispatched staff members (0.08%) was infected during the period while she was working at the front line under level 2 protection. The incidence of infection among healthcare workers with level 2 protection (0.08%, 1/1286) was lower than that observed among workers not using level 2 protection (2.9%, 119/4036). Statistical analysis revealed that the medical staff members who took inadequate protection had a higher risk of contracting infection than those using level 2 protection (relative risk: 36.9; 95% confidence interval: 5.2–263.6; Table 2).

**Table 2.**
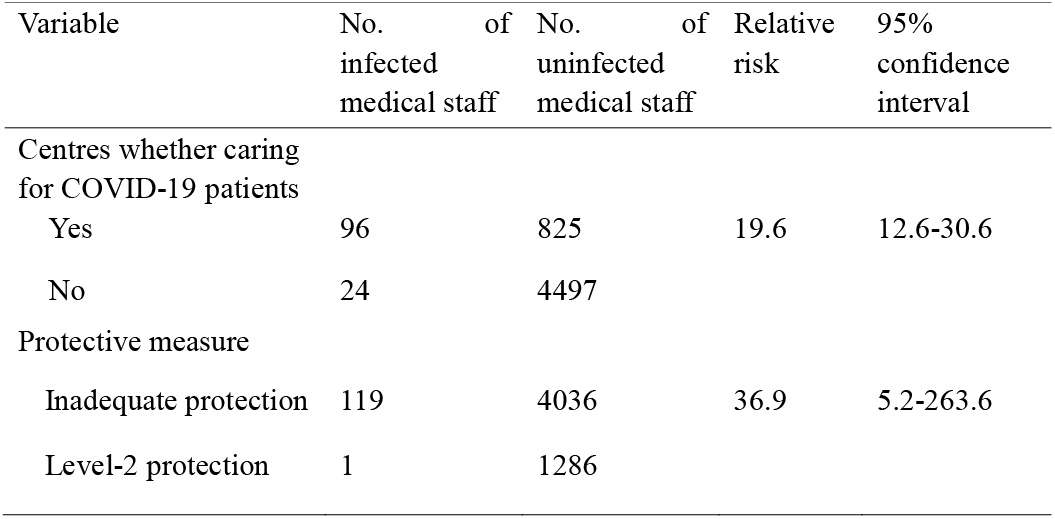
The relationship between infection and protective measure and whether COVID-19 patients treated

### The relationship between infection and COVID-19 patients treated

Neurosurgery departments in sixteen hospitals admitted a total of 59 patients with craniocerebral diseases complicated by laboratory-confirmed SARS-CoV-2 infection. Of the total number of infected staff members observed in this study, 96 (80.0%, 96/120) worked in these facilities, whereas 25 (20.0%, 25/120) worked in centres that did not receive any COVID-19 patients (Table 1). The prevalence of infection among the medical staff members in centres receiving COVID-19 patients (10.4%, 96/921) was higher than that of centres not receiving COVID-19 patients (0.5%, 24/4521). Statistical analysis revealed that medical staff members in centres receiving COVID-19 patients had a higher risk of contracting infection than those in centres not receiving COVID-19 patients (relative risk: 19.6; 95% confidence interval: 12.6–30.6; Table 2).

### Details of the doctor who died

In this case series, one medical worker died of COVID-19, and thereby, the fatality rate was 0.8%. He was a neurosurgeon above fifty in Wuhan. He began to develop fever and cough around 15 January 2020, but did not go to the hospital for examination. On 28 January, he was diagnosed with COVID-19 and thus hospitalised. Eventually, his condition deteriorated, and he died on 18 February.

## Discussion

This is the first report of the epidemiological characteristics of COVID-19 in affected medical staff members across several hospital centres in China. The survey included more than 80% of the centres with independent neurosurgical departments in Hubei province. In these neurosurgery departments (a medical field not specialised in the treatment of respiratory diseases), 121 cases of COVID-19 were observed among the medical staff. Aside from the 121 cases confirmed to be positive by the nucleic acid test, there were more than 300 symptomatic cases with or without positive radiological findings in this series. Due to the relatively low sensitivity of this particular nucleic acid test, it could not be excluded that there were several COVID-19-infected individuals also within this wider group of staff members. They all accepted home or hotel isolation. Therefore, the impact of COVID-19 among the neurosurgical healthcare professionals in Hubei province might have been even greater than our data indicate. The high number of cases among the medical staff members prompted us to analyse the causes of nosocomial infection and to provide a reference to the local authorities fighting the communicable disease.

This study investigated 5,443 medical staff members in neurosurgery departments in 107 hospitals. The overall incidence was 2.2%. Wuhan, the original epicentre of the outbreak, was the most affected area, accounting for 80% of all cases (96/120). These series of infection among medical staff members mainly comprised non-severe cases (87.6%). There were 119 (99.2%) patients who have already recovered and have been discharged. The fatality rate of 0.8% observed in the present study was similar to that of a recent large sample study.^20^

In daily work, medical staff members in neurosurgery departments usually only wear masks, hats and gloves when performing operations. A fairly large number of staff members do not take protective measures during ward rounds and outpatient visits. Unlike an isolation ward, neurosurgical wards have a highly mobile population.

Once an infectious disease emerges, it can lead to multiple infections: as an example, we present a typical triad of patient–medical staff–medical staff infection. On 25 December 2019, a patient with pituitary adenoma was hospitalised for surgery in the Department of Neurosurgery, Wuhan Union Hospital. Prior to hospitalisation, the patient exhibited no COVID-19 symptoms. The patient developed fever following surgery and was diagnosed with COVID-19 on 19 January 2020. Four nurses who had direct contact with the patient subsequently became infected. A number of medical staff members who had contact with the patients also developed fever, cough and other symptoms, of whom eight were confirmed to be infected with COVID-19 by laboratory testing. The presumed transmission tree is presented in Figure 4. Similar outbreaks occurred at three other centres, including the centre with the largest number of infections, i.e., 28. These findings agree with the report on the outbreak of a family cluster. ^10^

**Figure 4.**
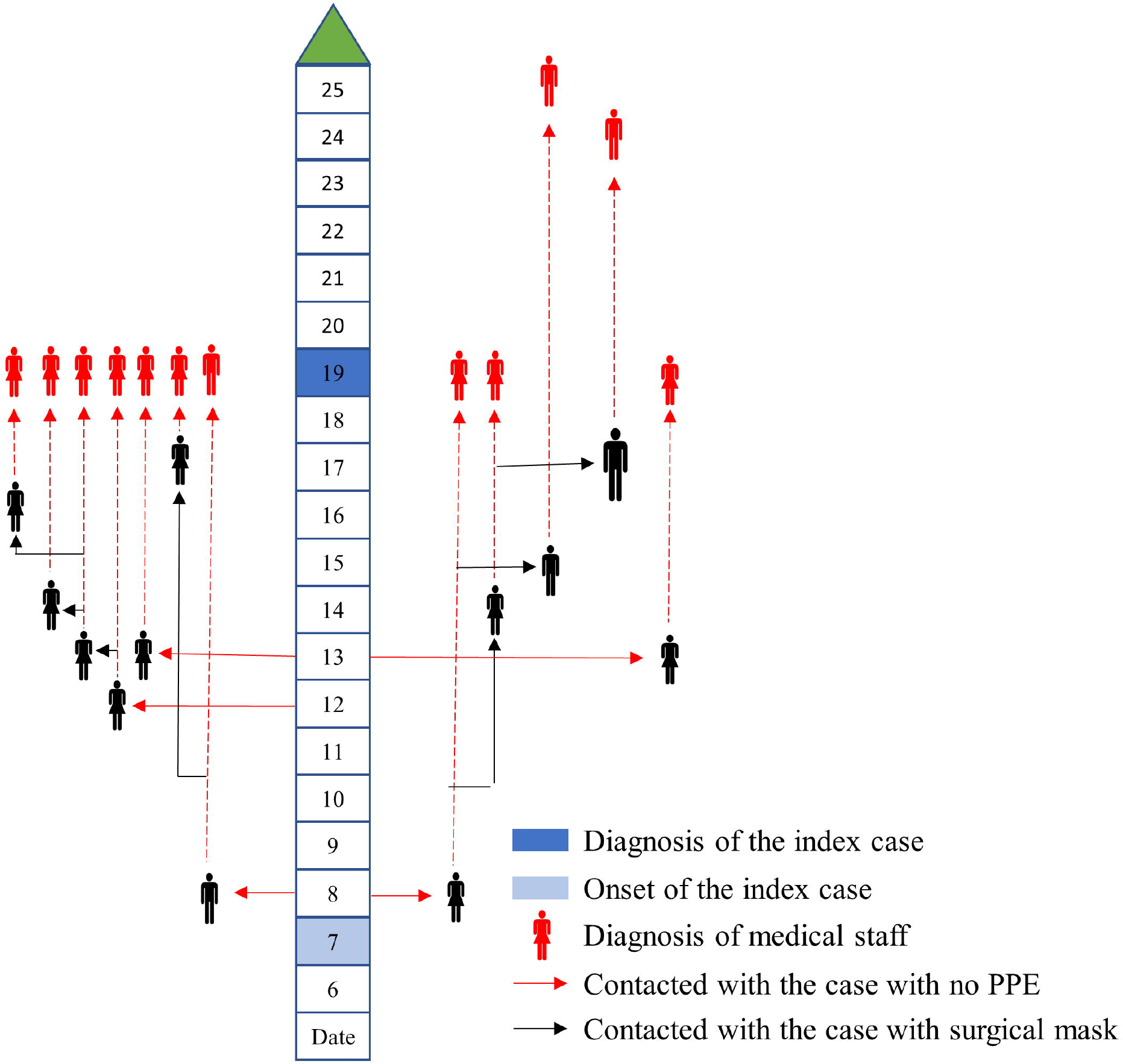
The presumed transmission trees of a COVID-19 patient to 12 medical staff members. The start date was 6 January 2020. PPE: personal protective equipment.

On 20 January, the National Health Commission of the People’s Republic of China classified the COVID-19 as a class b infectious disease and adopted control measures for class a infectious diseases. Since then, the medical organisations’ protective measures against COVID-19 have been strengthened. Many of the centres in this study began rigorous screening of patients prior to admission. Some centres converted inpatient wards into isolation wards. At the same time, most of the medical staff members adopted level 2 protection at work, and suspected cases were already treated in isolation. Once confirmed, they were immediately transferred to a special hospital for isolation treatment. In this series, 112 medical staff members (93.3%) were infected by 1 February. However, from 9 February to the end of the follow-up period, no more medical staff members were infected. This indicates the importance of protective measures in preventing infection.

We found that there was a close relationship between medical staff infection and whether COVID-19 patients were admitted to the department. In the 16 centres that treated COVID-19 patients, there were a total of 96 medical staff infections, accounting for 80% of all infections. However, in the 91 centres that did not treat COVID-19 patients, only 24 cases of infections occurred. Statistical analysis revealed that the risk of contracting infection among medical staff members in centres receiving COVID-19 patients was 19.6 times than those in centres not receiving COVID-19 patients. They were infected by patients or by those medical staff members who were infected during outpatient visits or consultations. One case contracted infection from family members. We could not rule out the possibility of other community infections in this series. However, from the above data, we could infer that most medical staff members were infected by patients directly, or indirectly by medical staff members who were infected by patients. On the one hand, compared with infectious disease specialist staff, neurosurgical specialist staff members are less aware of the protection against infectious diseases in their daily work. On the other hand, in the early days of the COVID-19 outbreak, the disease was poorly understood. The diagnosis of this disease was difficult due to the diversity of symptoms and a lack of nucleic acid testing. Inadequate protection against infection led to outbreaks among healthcare workers.

Our study revealed that protective measures play a significant role in preventing the spread of SARS-CoV-2. Most SARS-CoV-2 infections occurred in the initial stages of the outbreak, when the healthcare workers took only general or no protection measures. Later on, and with the improvement of protective measures, the incidence of infection decreased. A total of 1,287 medical personnel had been deployed to participate in the frontline response to COVID-19. However, only one healthcare worker was infected using level 2 protection. Statistical analysis revealed that the risk of contracting infection among medical staff members who took inadequate protection was 36.9 times than those who used level 2 protection. According to a report released by the National Health Commission of the People’s Republic of China, more than 42,000 medical workers from other provinces were dispatched to Hubei province, specifically to manage COVID-19. They fought on the front line using level 2 protection and were isolated in specific hotels at rest. However, none of them became infected (http://www.nhc.gov.cn/), indicating that proper protection measures could effectively prevent SARS-CoV-2 transmission.

Being based on neurosurgical staff members, this investigation reflects the microcosm of non-frontline departments in hospitals in Hubei province, China, the area most severely affected by COVID-19. The virus has spread into other continents, including Northern America, Southern Asia and Europe.^21-23^ With the strong intervention of the government, the outbreak in China has been well controlled. Our findings provide important reference information for medical facilities in all the areas where the disease is on the rise. First, COVID-19 is a highly contagious infectious disease. However, proper protective measures can effectively prevent medical workers from being infected. The results of this study suggest that level 2 or a higher level of protection should be used when managing COVID-19. Even in hospital units not directly treating COVID-19 patients, protection awareness and measures should not be neglected. In the end, once suspected cases are identified, strong quarantine measures on those in close contact with the source of infection should be taken to prevent the spread of infection. In view of China’s successful experience in fighting the epidemic, strong control and social distancing policies may have played significant roles in reducing the rate of virus transmission.

## Limitations of this study

Our study has limitations. Firstly, due to the large number of centres involved, the variables of the study are relatively simple, and the results do not cover all the epidemiological characteristics of the neurosurgical medical workers in Hubei province infected with SARS-CoV-2. Secondly, identifying the source of infection was difficult, and our extrapolations might not be accurate. Thirdly, the protective measures adopted by medical staff members were not fixed but changed over time. Therefore, the analysis based on protective measures might be affected by time bias. Fourthly, the self-reported questionnaire survey was prone to responder bias. The respondents’ descriptions might be inconsistent with the facts, which could affect the reliability of the results. Lastly, some cases had uncertain documentation of the exposure history, and recall bias might exist in the epidemiological investigation.

## Conclusion

In conclusion, medical staff members in the neurosurgical hospital units in Hubei province were seriously affected by COVID-19. Most of the infections were attributable to inadequate precautions taken by medical staff members in the early stages of the outbreak. Level 2 protection is effective in preventing infection among medical workers. The enforcement of rigorous protection is also of great importance in hospital units that are not directly involved in combatting COVID-19.

## Data Availability

The corresponding author could provide data if requested.

## Author Contributions

### Concept and design

Jincao Chen, Qianxue Chen, Hongyang Zhao.

### Acquisition, analysis, or interpretation of data

Yansen Bai, Xuan Wang, Haijun Wang, Xuebin Hu, Feng Wang, Xianke Wang.

### Drafting of the manuscript

Qiangping Wang, Xing Huang.

### Critical revision of the manuscript for important intellectual content

Jincao Chen, Qianxue Chen, Xiaobing Jiang, Hongyang Zhao.

### Statistical analysis

Qiangping Wang, Xing Huang, Yansen Bai.

### Administrative, technical, or material support

Xuan Wang, Haijun Wang, Xuebin Hu, Feng Wang, Xianke Wang, Xiaobing Jiang.

### Supervision

Jincao Chen, Qianxue Chen, Xiaobing Jiang, Hongyang Zhao.

## Acknowledgements

We thank the directors of each medical centre for allowing us to collect the data. We offer our most sincere condolences to the neurosurgeon who has dedicated his live to fighting disease.

